# How Much Does the Reduced EEG Montage Matter for Seizure Detection?: A Large-Cohort Simulation Study

**DOI:** 10.64898/2026.05.05.26352477

**Authors:** Joe Kojima, Haoer Shi, Svanik Jaikumar, William K.S. Ojemann, Carlos Aguila, Juri Kim, Taneeta Mindy Ganguly, Brian Litt, Erin C. Conrad

## Abstract

**Importance:** Implantable sub-scalp EEG systems with a small number of channels have emerged as promising solutions for long-term seizure monitoring in patients with epilepsy. How seizure detection performance varies by montage configuration is unknown.

**Objective:** To quantify how automated seizure detection performance differs between full and reduced montages, and how these differences vary by epilepsy characteristics.

**Design:** Retrospective cross-sectional study.

**Setting:** Single-center at the Hospital of the University of Pennsylvania Epilepsy Monitoring Unit (EMU).

**Participants:** EEG data from 2281 consecutive EMU admissions between January 2017 and December 2024 were screened. Admissions with at least one annotated seizure and one interictal clip ≥20 minutes from any seizure were included.

**Exposure:** Computational simulation of published sub-scalp device montages using standard 10-20 EEG channels.

**Main Outcomes and Measures:** The primary outcome was event-based F1 scores evaluated for three published seizure detectors—a one-class support vector machine (SVM), a convolutional neural network (SPaRCNet), and a long short-term memory autoregressive model (NDD)—across montages.

**Results:** A total of 466 admissions from 436 patients (mean [SD] age, 39.0 [14.4] years; 54.4% female) met inclusion criteria, comprising 1683 seizures and 1527 interictal clips. SPaRCNet achieved the highest performance (mean [SD] F1, 0.61 [0.30]), followed by NDD (0.56 [0.28]) and SVM (0.39 [0.25]). Performance decreased by at most 0.09 with reduced montages, depending on detectors. Patient factors accounted for the largest proportion of performance variance (29.2%), followed by detector choice (10.3%). Montage effects were minimal (0.4%), despite variation in optimal montage across detectors. Reduced-montage performance correlated moderately to highly with full-montage performance (ρ=0.29–0.73), suggesting full-montage performance could help identify patients suitable for sub-scalp devices. Missed seizures were associated with lower amplitude and bandpowers than detected seizures, though they remained distinguishable from interictal data.

**Conclusions and Relevance:** Automated seizure detection achieved comparable accuracy, with only modest reductions, under simulated reduced montages. Performance differences were driven primarily by detector- and patient-level factors rather than montage. These findings support the feasibility of accurately detecting seizures with published sub-scalp devices and highlight the need for improved algorithms to optimize performance.

**Key Findings:** *Question:* How do automated seizure detection algorithms perform with reduced-channel montages simulating published sub-scalp devices?

*Findings:* In this retrospective cross-sectional study, seizure detection performance decreased only modestly on reduced montages relative to the full montage (absolute F1 change −0.09 to 0.014), whereas patient- and algorithm-level factors accounted for most of performance variance (29.2% and 10.3%, respectively). Algorithm performance on full montage recordings was moderately correlated with performance on reduced channel montages (ρ=0.29–0.73).

*Meaning:* Reduced-montage sub-scalp devices are promising for ultra-long-term monitoring, but best performance requires selecting the right patients. Patient-specific seizure detectors will likely be required to optimize long-term performance.

## Introduction

Accurately counting seizures is critical for guiding therapy in drug-resistant epilepsy (DRE) patients. However, current clinical practice relies on patient self-reporting, which is notoriously inaccurate. Patients remain unaware of over half of their seizures, particularly those occurring during sleep or manifesting as subtle impaired awareness events ^1^. Consequently, objective electrophysiological monitoring remains central to effective disease management.

Inpatient Epilepsy Monitoring Unit (EMU) monitors seizures by recording continuous EEG in a controlled setting. However, the short duration of EMU admissions precludes assessing long-term temporal dynamics ^2^. Ambulatory EEG enables at-home recording with similar electrode coverage, but is typically limited to several days due to discomfort and electrode detachment ^3^.

Ultra-long-term, ambulatory monitoring is poised to bridge this gap ^4–7^. Recent advances enable minimally invasive, sub-scalp implantable devices for chronic use, which record EEG via a small number of electrodes typically placed in the subgaleal or subcutaneous space. These systems make continuous, multi-year monitoring possible, providing objective outpatient EEG data to inform epilepsy management. It is unknown how transitioning from standard, full-scalp electrode coverage to low-density devices impacts seizure detection accuracy.

The scale of data generated by long term monitoring devices makes manual review impractical, underscoring the need for reliable automated seizure detection methods. Machine learning (ML) algorithms have shown strong seizure-detection performance using full-density EEG montages ^8–11^. Recent studies have also demonstrated the feasibility of automated seizure detection for sub-scalp devices using an algorithm tailored to the device configuration, though performance has been variable ^6,12–14^. It remains unclear whether models trained on full-scalp EEG are robust to reduced-channel data, or how performance varies across montages. Understanding how seizure detection performance depends on reduced EEG montage designs and epilepsy localization is essential to guiding which patients are candidates for these devices and where they are placed.

In this study, we evaluate the performance of multiple validated automated seizure detectors on computationally reduced EEG montages simulating configurations of published sub-scalp devices. We benchmark seizure detection performance against full-montage baselines, quantify changes in performance across reduced montages likely to be applied in clinical settings, and explore their dependence on epilepsy classification, lateralization, and localization. Our findings provide a computational assessment of seizure detection performance using reduced-channel montages and highlight considerations for their future clinical application.

## Methods

### Study cohort

This was a single-center retrospective study of data from 2281 consecutive admissions at the Hospital of the University of Pennsylvania (HUP) Epilepsy Monitoring Unit (EMU) between January 2017 and December 2024. The study protocol (#835008) was approved by the Institutional Review Board (IRB) of the University of Pennsylvania. Inclusion criteria were: 1) EEG data available on the cloud clinical server, 2) At least one epileptic seizure with clearly annotated electrographic onset and offset as determined by the clinical treatment team, and 3) At least one interictal clip available from the same admission (**eFigure 1**). EEG data were acquired at 256 Hz, with electrodes placed according to the international 10–20 system (see **eMethods**). Demographic information and epilepsy characteristics were extracted from clinical notes, including age, sex, epilepsy classification (focal, generalized, mixed, unknown), lateralization (left, right, bilateral, unknown), and localization (temporal, frontal, multifocal, unknown).

### Signal preprocessing

Seizure clips were extracted including a 5-minute padding before seizure onset and after offset. For each admission, 10-minute interictal clips, matched in number to the seizure clips, were sampled ≥20 minutes from any seizure-related annotation, preferably from the same day as seizures. Fewer interictal clips were included when insufficient data were available (see **eMethods**). EEG signals were bandpass filtered between 0.5-100 Hz and notch-filtered at 60 Hz.

### Montage configurations

Full montage data were re-referenced to 16 standard bipolar pairs (**eTable 1**). To simulate signals from existing sub-scalp long-term EEG monitoring devices, we constructed bipolar pairs with electrodes in the 10–20 system nearest to reported implant locations. We chose to primarily simulate montages corresponding to two devices—the Minder system (Epiminder, Melbourne, Australia) and the 24/7 SubQ implant (UNEEG Medical, Lillerød, Denmark)—chosen because, to our knowledge, these devices are the only chronic implanted EEG recording devices that have FDA approval or CE marking, as well as the only ones with published clinical trials in the ambulatory setting ^5–7,15^. We simulated the Minder design using centroparietal channels C3–P3 and C4–P4, and the most commonly described configuration in the original UNEEG trial ^6^ using frontotemporal electrode pairs F4–T4 (right-sided implant), F3–T3 (left-sided implant), and both (bitemporal implant) (**Figure 1B**). Additional configurations, including a circumferential “hatband” montage, were simulated as supplemental analyses (**eTable 1**).

**Figure 1.**
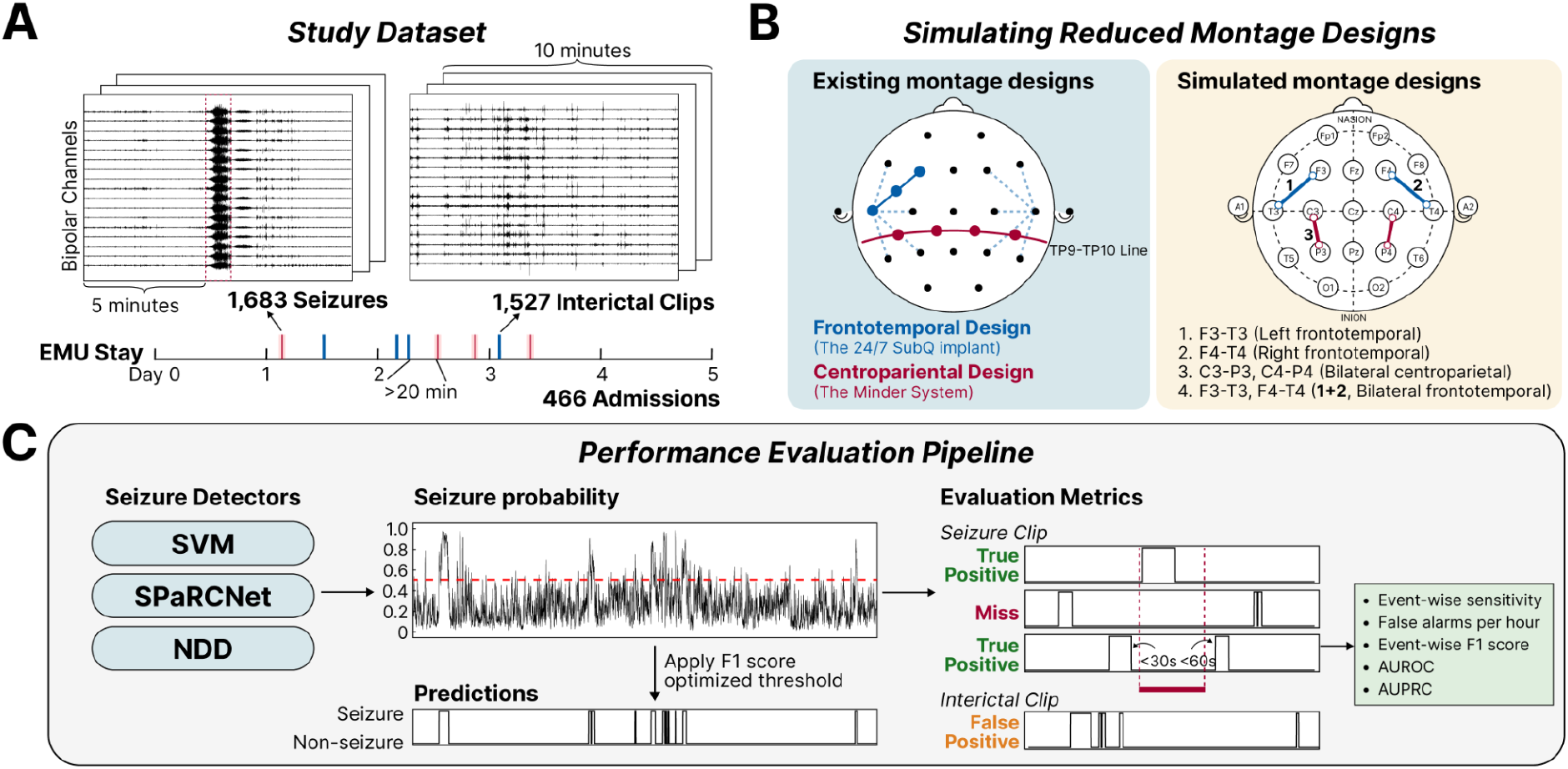
Methods overview. **A. Study Dataset**. A total of 1,683 seizure clips and 1,527 10-minute interictal clips were sampled from 466 EMU admissions. Clinician-confirmed seizures were extracted with 5-minute padding before and after each event. For each admission, we attempted to extract an equal number of interictal clips from periods at least 20 minutes away from any suspected seizure activity and, when possible, from the same day as seizures. **B. Subscalp reduced montages simulated**. Two existing sub-scalp reduced montage designs - the frontotemporal design from the 24/7 SubQ implant (UNEEG Medical) and the centroparietal design from the Minder System (Epiminder) - were simulated using bipolar pairs from the 10–20 system (yellow box). The Minder System implant consists of four recording contacts along the TP9–TP10 line of the standard 10–10 system, forming one bipolar pair over each hemisphere (blue box, red line). The 24/7 SubQ implant consists of three electrodes on a single lead, centered at the postauricular region and oriented toward the region of interest (blue box, blue lines). Two bipolar channels were formed using the distal and proximal electrodes. Depending on the patient’s seizure laterality, one or two leads could be implanted on the left, right, or bilaterally. The most commonly used orientation for the SubQ implant was used for simulation. **C. Performance evaluation pipeline**. The performance of 3 different automated seizure detectors (SVM, SPaRCNet, and NDD) was evaluated on the full and reduced montage designs. For each detector, a threshold that optimized F1 score across the whole dataset was applied to obtain binary predictions. Consecutive seizure predictions were grouped into events. Event-wise sensitivity was defined as the percentage of seizures detected by the algorithm, where a prediction overlapped with the event, allowing a 30-second pre-ictal and 60-second post-ictal tolerance. The false alarm rate per hour was defined as the number of false predictions that did not overlap with any true seizure events, normalized by recording duration. We additionally assessed event-wise F1 score, AUROC (area under the receiver operating characteristics curve), and AUPRC (area under the precision-recall curve).

### Seizure detectors

The impact of reduced montages on seizure detection performance may depend on the detection algorithm. To isolate the effect of montage reduction, we evaluated multiple seizure detection algorithms representing diverse, validated approaches, including a simple one-class support-vector machine (SVM) that learns the feature patterns ^9^, a deep convolutional neural network pretrained on a large volume of scalp EEG data (SPaRCNet) ^10^, and a long short-term memory (LSTM) model measuring neural dynamic divergence from patient-specific baseline (NDD) ^11^. See **eMethods** for details.

### Performance evaluation

EEG signals, in full or reduced montages, were input to each detector to generate seizure probabilities in sliding windows of 1-10 seconds, depending on the detector. As different montages led to shifts in predicted probability distributions, decision thresholds were optimized separately for each montage to maximize event-wise F1 score across all patients. These thresholds were applied to raw probabilities to generate binary seizure predictions, which were then smoothed to keep only consecutive positive (seizure) predictions ≥20 seconds, defined as seizure events (see **eMethods**).

We used the SzCORE framework to evaluate seizure detection performance ^16^. An annotated seizure was considered detected if any prediction overlapped with it, allowing 30-second pre-ictal and a 60-second post-ictal tolerance (**Figure 1C**). Predicted events overlapping with seizures were counted as true positives; otherwise, as false positives. Unmatched annotated seizures were counted as misses. Metrics were computed at the event level, including (1) event-wise sensitivity, defined as percentage of annotated seizures being detected; (2) false alarm rate, defined as number of false positives per hour; (3) event-wise F1 score, computed from (1) and event-wise precision (percentage of predicted events being true positives), which was used as the primary metric. Metrics were aggregated across all clips within each admission. We further evaluated stratified performance by patient epilepsy classification, lateralization, and localization.

### Missed seizure characteristics

To investigate characteristics of missed seizures, we extracted a set of univariate time-series and spectral features, including amplitude, envelope, variance, line length, and bandpower across six canonical frequency bands for interictal clips and seizures (**eMethods**). Missed versus detected status was defined based on full montage results of SPaRCNet model, given its overall highest performance. To identify features associated with missed seizures, we fitted a logistic regression model with missed status as the outcome and features as predictors. Feature importance in prediction was estimated using a permutation method (**eMethods**) ^17^. Finally, as proof-of-concept to assess whether missed seizures could potentially be detected by a future detector, we trained a logistic regression model to predict ictal status using features in 5-fold cross-validation. Receiver operating characteristics (ROC) and precision–recall (PR) curves were generated from model-predicted probabilities, separately for detected and missed seizures.

### Statistical Analysis

Performance across montages was compared using the Friedman test for repeated measures. When significant, post hoc pairwise comparisons were performed using Wilcoxon signed-rank tests, with effect sizes calculated as *r* = |*Z*| / √*N*. A Mann–Whitney U test was used to compare feature differences between groups. P values were adjusted for multiple comparisons using the Benjamini–Hochberg procedure, with statistical significance defined as a 2-sided adjusted *p*<0.05. We additionally used linear mixed effects models to 1) measure the proportion of variance explained by patients, detector, and montage, and 2) evaluated the influence of epilepsy classification, laterality, and localization on performance (**eMethods**).

## Results

### Study cohort and epilepsy characteristics

The final cohort included 466 admissions from 436 unique patients, yielding 1,683 clinician-confirmed seizures and 1,527 interictal clips (**Figure 1A**). The study population had a mean (SD) age of 39.0 (14.4) years at first admission and was 54.4% female (**Table 1**). The median number of seizures per admission was 2.0 (IQR, 1.0–4.0), with a mean (SD) seizure duration of 81.4 (70.8) seconds. Most patients had focal epilepsy (75.9%), followed by generalized (9.9%), and mixed (6.2%) classifications. Among patients with focal or mixed epilepsy (n=358), 43.6% had left-sided epilepsy, 28.5% right-sided, 16.2% bilateral, and 11.7% unknown; seizure localization was temporal in 67.0%, frontal in 6.2%, multifocal in 10.1%, and unknown in 16.8%.

**Table 1.** Patient demographics and seizure characteristics.

| **Variables** | **Overall (n=436)** |
| --- | --- |
| Age at 1st admission, mean (SD) | 39.0 (14.4) |
| Sex, n (%) |  |
| Female | 237 (54.4) |
| Number of Seizures, median [Q1,Q3] | 2.0 [1.0,4.0] |
| Seizure Duration (sec), mean (SD) | 81.4 (70.8) |
| Epilepsy Classification, n (%) |  |
| Focal | 331 (75.9) |
| Generalized | 43 (9.9) |
| Mixed | 27 (6.2) |
| Unknown | 35 (8.0) |
| Epilepsy Lateralization for focal/mixed patients, n (%), n=358 |  |
| Left | 156 (43.6) |
| Right | 102 (28.5) |
| Bilateral | 58 (16.2) |
| Unknown | 42 (11.7) |
| Epilepsy Localization for focal/mixed patients, n (%), n=358 |  |
| Temporal | 240 (67.0) |
| Frontal | 22 (6.2) |
| Multifocal | 36 (10.1) |
| Unknown | 60 (16.8) |

### Seizure detection performance across detectors and montages

We evaluated the seizure detection performance of three detectors—SPaRCNet, NDD, and SVM—across the full and simulated reduced-channel configurations. Performance varied substantially across detectors and patients. In a linear mixed-effects model, performance depended on detector (*χ*^2^(2)=166.90; *p*<.001), and the detector–montage interaction (*χ*^2^(8)=63.20; *p*<.001), with no significant effect of montage after accounting for these effects (*χ*^2^(4)=1.95; *p*=.75). Patient admission accounted for the largest proportion of variance in performance (29.2%), followed by detector choice (10.3%), with minimal contributions from montage (0.4%) and the montage–detector interaction (0.5%) (**Figure 2A, eTable 2**). Overall, seizure detection accuracy strongly depended on admission and on the automated detection algorithm, with only a minor effect of montage.

**Figure 2.**
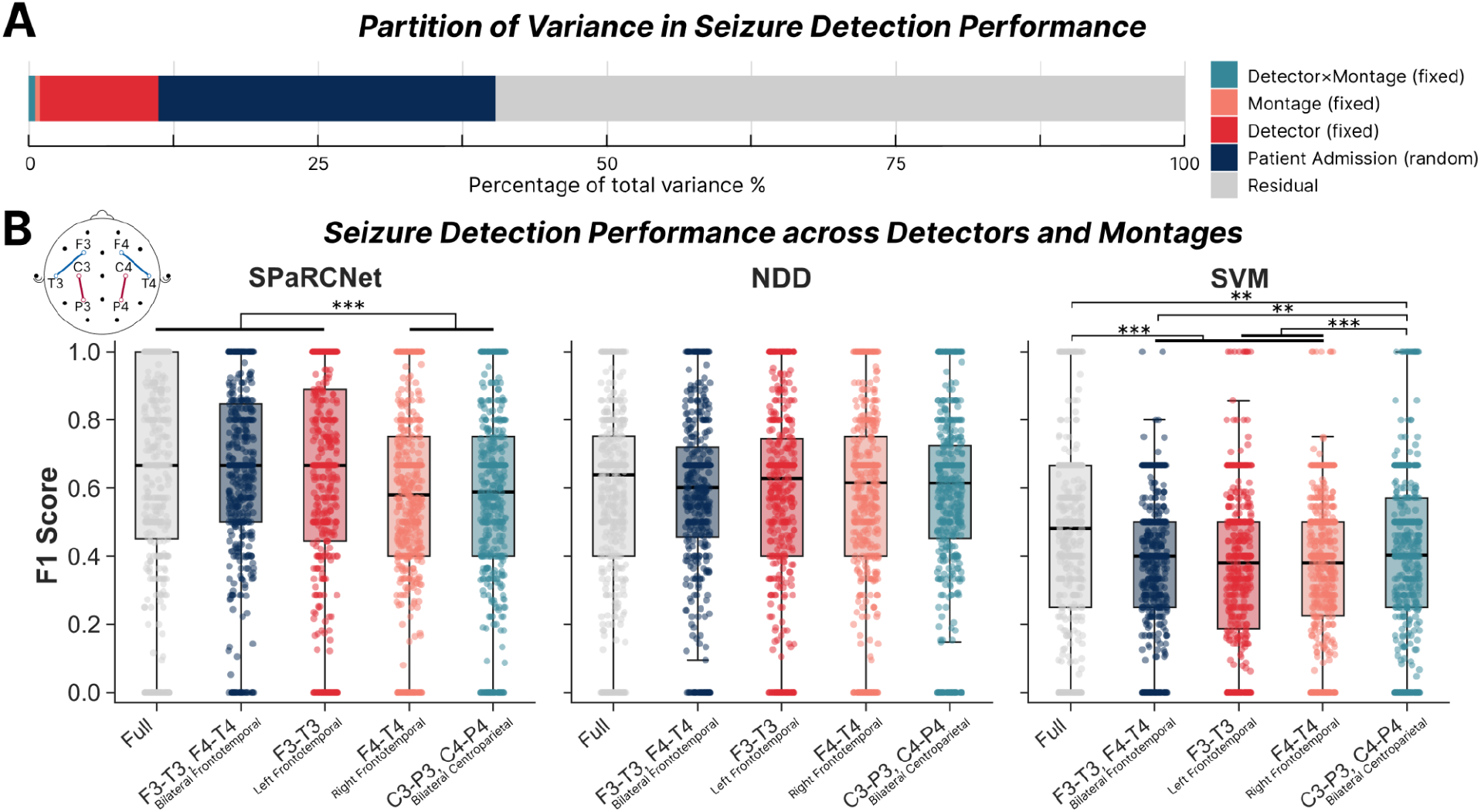
Seizure detection performance. **A. Partition of variance in seizure detection performance**. We built a linear mixed-effects model with seizure detection F1 score as outcome, seizure detector, montage, and their interaction as fixed effects predictors, and patient admission as random intercept. There was 59.6% residual variance, representing unexplained error. The random effects accounted for 29.2% of the total variance, with detector, montage, and their interaction contributing 10.3%, 0.4%, and 0.5%, respectively. This suggests that seizure detection performance is highly dependent on the specific patient admission and partly dependent on the choice of seizure detector, with minimal effect of the montage or the detector-montage interaction. **B. Seizure detection performance across detectors and montages**. F1 scores for the SPaRCNet, NDD, and SVM detectors are shown across montages. Each dot represents one EMU admission. Boxes indicate the interquartile range (IQR), with the central line representing the median across admissions; whiskers extend to 1.5×IQR. A Friedman’s test was used to assess overall differences across montages, followed by post hoc Wilcoxon signed-rank tests for pairwise comparisons. P-values were adjusted for multiple comparisons using the Benjamini-Hochberg method. SPaRCNet, Friedman’s test *χ*^2^(4)=78.60, *p*<.001; Wilcoxon signed-rank test full vs. F4-T4, W=52188, adj. *p*<.001, *r*=0.25; full vs. C3-P3,C4-P4, W=52274, adj. *p*<.001, *r*=0.27; F3-T3,F4-T4 vs. F4-T4, W=44601, adj. *p*<.001, *r*=0.38; F3-T3,F4-T4 vs. C3-P3,C4-P4, W=51,475.5, adj. *p*<.001, *r*=0.27; F3-T3 vs. F4-T4, W=46386, adj. *p*<.001, *r*=0.21; F3-T3 vs. C3-P3,C4-P4, W=50161, adj. *p*<.001, *r*=0.20. NDD, Friedman’s test *χ*^2^(4)=6.51, *p*=.16. SVM, Friedman’s test *χ*^2^(4)=75.78, *p*<.001; full vs. F3-T3,F4-T4, W=54343.5, adj. p<.001, r=0.28; full vs. F3-T3, W=53562, adj. *p<*.001, *r*=0.29; full vs. F4-T4, W=53950.5, adj. *p*<.001, *r*=0.30; full vs. C3-P3,C4-P4, W=48429.5, adj. *p*<.001, *r*=0.16; F3-T3,F4-T4 vs. C3-P3,C4-P4, W=26721, adj. *p*=.002, *r*=0.17; F3-T3 vs. C3-P3,C4-P4, W=25152.5, adj. *p*<.001, *r* = 0.18; F4-T4 vs. C3-P3,C4-P4, W=25719.5, adj. *p*<.001, *r*=0.20. ***, *p* < .001; **, *p* < .01; *, *p* < .05.

SPaRCNet achieved the highest mean F1 score across montages (0.607 [SD, 0.30]; sensitivity 0.788 [SD, 0.34]; false alarm rate 2.72 alarms/hour [SD, 2.89]), followed by NDD (0.564 [SD, 0.28]; 0.785 [SD, 0.35]; 3.08 [SD, 2.42]), and the SVM model (0.391 [SD, 0.25]; 0.668 [SD, 0.37]; 5.86 [SD, 2.77]). For SPaRCNet and SVM, performance differed significantly across montages (Friedman test: SPaRCNet, *χ*^2^(4)=78.60, *p*<.001; NDD, *χ*^2^(4)=6.52, *p*=.16; SVM, *χ*^2^(4)=75.78, *p*<.001). Post hoc comparisons demonstrated detector-specific patterns (**Figure 2B**). Despite this variability across montages, performance decreases under reduced montages were small, ranging from 0.005-0.08 for SPaRCNet, −0.014 to −0.001 for NDD, and 0.048-0.09 for SVM. Sensitivity, false alarm rate, and ROC and PRC curves showed similar trends (**eTables 3-5, eFigure 2**), noting that the sensitivity at the F1-optimized decision threshold paradoxically *increased* for some simulated reduced montages, corresponding to an increase in the false alarm rate. Spearman’s correlation showed strong concordance of patients’ performance between full and reduced montages for NDD (ρ=0.63–0.73) and weaker concordance for SPaRCNet (ρ=0.30–0.43) and SVM (ρ=0.29–0.33) (**eFigure 3**). We restricted subsequent analyses to the SPaRCNet and NDD detectors to limit the number of comparisons, given the poor performance of the SVM detector. Results for other simulated montages are available in **eTables 3-5**.

### Performance Variations by Epilepsy Characteristics

We tested how detector performance varied with epilepsy characteristics (**Figure 3, eTable 6**). For SPaRCNet, seizure detection performance was independent of epilepsy classification (*χ*^2^(1)=1.237, *p*=.27) and its interaction with montage (*χ*^2^(4) = 3.288, *p*=.51). In contrast, NDD showed a significant effect of classification (*χ*^2^(1)=5.001, *p*=.025), with higher accuracy in generalized compared to focal epilepsy. The classification-montage interaction was not significant (*χ*^2^(4)=8.663, *p*=.07).

**Figure 3.**
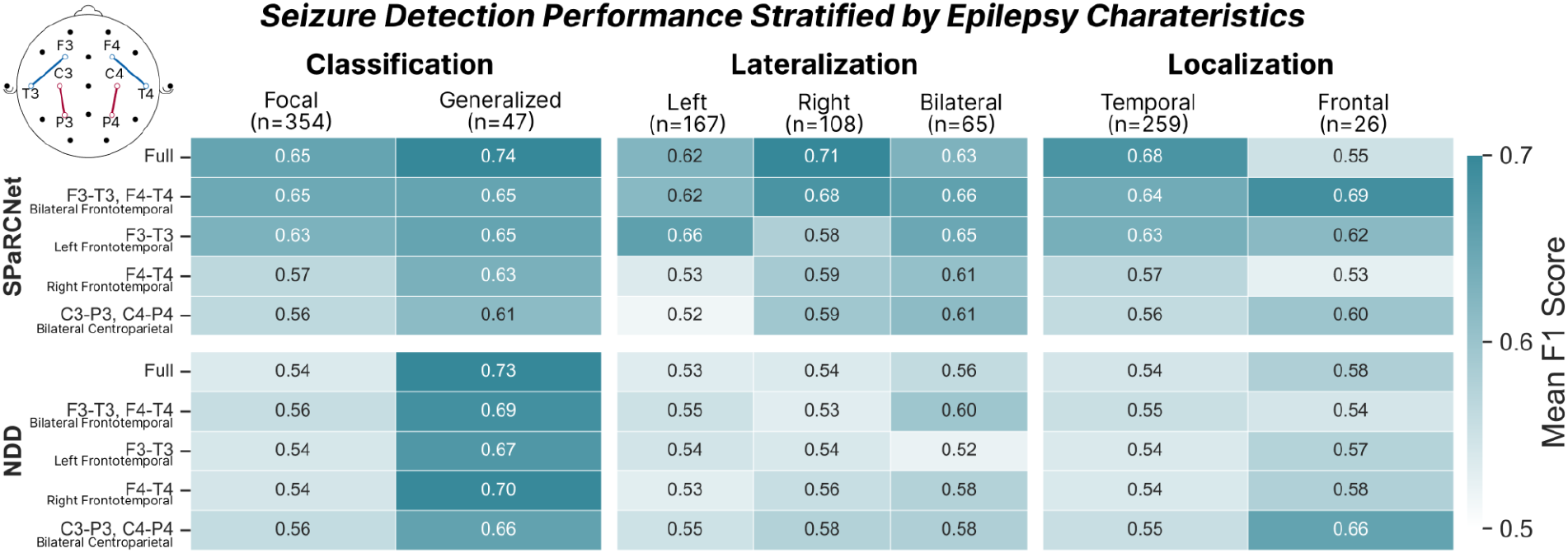
Seizure detection performance across clinical subgroups. Mean event-wise F1-scores are shown for SPaRCNet (top) and NDD (bottom) across different electrode montage configurations. Performance is stratified by epilepsy classification (focal, generalized), laterality (left, right, bilateral), and seizure location (temporal, frontal). Each cell represents the mean F1-score across EMU admissions within the corresponding subgroup. Sample sizes were indicated in parentheses. White-to-blue color scale reflects lower to higher performance.

For epilepsy lateralization, SPaRCNet showed significant effects of laterality and its interaction with montage (laterality: *χ*^2^(2)=8.098, *p*=.017; interaction: *χ*^2^(8)=30.21, *p*<.001). Patients with left-sided epilepsy had lower performance than those with right or bilateral epilepsy in all montages except for the left frontotemporal montage. In contrast, NDD showed no significant performance differences across laterality groups (*χ*^2^(2)=0.956, *p*=.62) or across montages (*χ*^2^(4)=9.229, *p*=.06). Epilepsy localization and its interaction with montage had no significant effects for either detector. Results for all other simulated reduced montage configurations are available in **eFigure 4**.

### Characteristics of missed seizures

Analysis of missed seizures indicated that they showed lower amplitude, variance, line length, bandpower, and shorter duration compared to detected seizures (*p*<.001, **Figure 4A**), despite features still being higher than in interictal clips (*p*<.001). Line length (0.180 [SE, 0.025]), theta power (0.090 [SE, 0.036]), and alpha power (0.086 [SE, 0.015]) showed highest permutation importance in predicting if a seizure was missed. A simple logistic regression model using these features yielded an AUROC of 0.710 and AUPRC of 0.051 in detecting windows of missed seizure from interictal, compared to 0.859 and 0.378, respectively, for detected seizures (**Figure 4B**).

**Figure 4.**
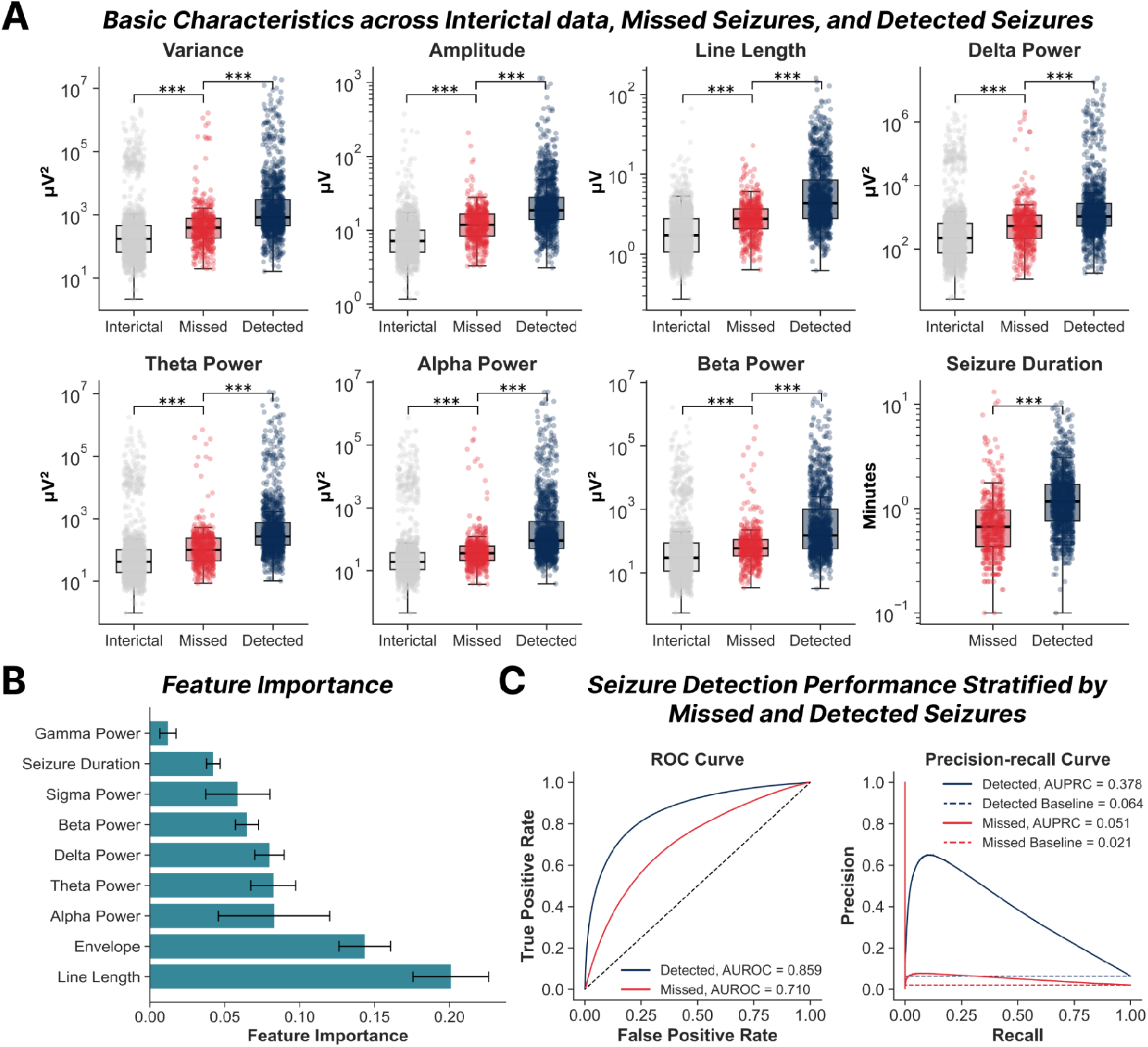
Characteristics of missed seizures. **A. Basic EEG characteristics across interictal clips, missed seizures, and detected seizures.** Data were calculated in non-overlapping 5-second windows channel-wise, then averaged across 16 standard 10-20 bipolar channels. For interictal clips, features from windows were averaged across full duration; for seizure clips, window-level features were averaged from seizure onset to offset. Each dot represents one clip or seizure. Mann–Whitney U tests were used to compare differences between interictal and missed seizures, and between missed and detected seizures, with p-values corrected using the Benjamini-Hochberg method. ***, p < 0.001. **B. Feature importance for predicting missed versus detected seizures**. Importance was computed as the permutation importance in 5-fold cross-validation. **C. Seizure detection performance stratified by missed and detected seizures**. Seizure probabilities were obtained through fitting a logistic regression model with seizure label as the outcome and log-transformed, standardized basic characteristics as predictors, using 5-fold stratified cross-validation at the patient admission level. ROC, receiver-operating characteristics. A seizure was classified as “missed” or “detected” based on whether it was missed or detected by the SpaRCNet model run on the full 10-20 montage, with “missed” and “detected” defined in the Methods.

## Discussion

In this large, heterogenous EMU cohort, we evaluated the performance of multiple automated seizure detectors across full scalp EEG and simulated reduced montages approximating published sub-scalp recording devices. Performance was moderate to good for the best-performing detector and was largely preserved across reduced montages, supporting the feasibility of ultra-long-term monitoring with sub-scalp devices. Montage choice had a relatively small impact; instead, performance variability was driven primarily by patient heterogeneity and detector choice. Performance on full and reduced montages was moderately correlated, suggesting that pre-implantation EMU recordings may help guide patient selection. Missed seizures exhibited features closer to interictal activity than detected seizures, however the missed seizures remained distinguishable from interictal activity, indicating the need for further optimizing detection algorithms.

Minimally invasive sub-scalp EEG systems have emerged as a promising approach for long-term outpatient monitoring ^4,18^, with potential applications in seizure counting, differential diagnosis, lateralization, surgical planning, and early warning for rescue medication or caregiver notification. However, whether seizures can be reliably detected using automated methods on these reduced montages remains unclear.

Our findings demonstrate that automated detection algorithms perform comparably on full and reduced montages, with several key insights. First, performance is highly dependent on the choice of seizure detector. Among the tested algorithms, SPaRCNet achieved the best performance, potentially reflecting its training on scalp EEG, in contrast to other models developed primarily on intracranial recordings with distinct signal characteristics. While sensitivity and F1 scores remained robust across reduced montages, false alarm (FA) rates highlight a persistent challenge for ultra-long-term ambulatory monitoring. SPaRCNet produced approximately 1.5–3.6 false alarms per hour, corresponding to dozens per day in a chronic outpatient setting. While such rates are manageable for retrospective review through quick, bulk dismiss of artifacts, they limit the utility of devices for real-time alerting. Future implementations may benefit from threshold tuning (trading sensitivity for lower FA rates) and human-in-the-loop review to suppress these false alarms ^19,20^. Notably, models trained specifically on ambulatory EEG data may achieve better performance. Prior studies of the UNEEG SubQ device using EpiSight, an automated algorithm specifically trained on two-channel data, have demonstrated promising performance, with a median (IQR) sensitivity of 70.5% (48–94.6%) and a lower false alarm rate than observed in our study of 3.4/day (3–15.8/day) ^6,13,14^.

Second, the individual patient was the most significant determinant of detection performance. Both the EpiSight studies and our findings observed substantial variability across patients. This has important practical implications: patient selection will be critical for the clinical utility of sub-scalp monitoring devices. Future clinical trials should consider baseline detectability of a patient’s seizures on a full 10-20 montage in addition to diagnosis. Given the concordance of performance on the full montage and reduced montages, algorithm performance during EMU admissions could help predict the likely benefit of sub-scalp monitoring, enabling more informed implantation decisions before an invasive procedure. Additionally, generalized models and uniform thresholds across all patients, employed in the current study, are unlikely to be optimal. Clinical applications may benefit from patient-specific algorithms and thresholds.

Third, while montage impact was modest compared with patient- and detector-level factors, we observed clinically intuitive patterns. For instance, left- and right-sided montages perform better in ipsilateral epilepsy. Although not statistically significant, potentially due to small localization subgroups, centroparietal montage performed better in frontal-lobe epilepsy. These trends suggest that device selection and implant location may be tailored to epilepsy etiology and inform montage choice. Notably, seizure detection performance was largely maintained even when the montage was contralateral to the side of the epilepsy, which may reflect that contralateral effects, spread or volume conduction can be detected in unilateral onset seizures.

## Limitations

Several limitations merit consideration. Our computationally reduced montages imperfectly approximate implant locations of SubQ or Minder systems, whose physical hardware arrays differ substantially. Scalp EEG also cannot fully reproduce properties of true sub-scalp systems, including electrode-tissue interface, impedance profile, and mechanical artifact behavior, despite prior work suggesting similar signals between sub-scalp and overlying scalp electrodes ^7^. Furthermore, ambulatory settings introduce environmental and muscle noise absent in EMU settings, potentially inflating false alarm rates. We used clinician annotations as ground truth, which may introduce localization inaccuracies and bias toward more obvious seizures, potentially overestimating sensitivity for subtle seizures. We used a uniform, F1-optimized threshold across all patients, which approximated a leave-one-patient-out strategy given the large sample size ^11^. Although this may overestimate absolute performance, it should not affect comparisons across montages. Finally, while we examined several established detection algorithms, alternative approaches may perform differently and warrant future evaluation ^21^. Consequently, our results likely represent a conservative baseline rather than a direct projection of clinical device efficacy.

## Conclusions

In a large, heterogeneous cohort of epilepsy patients, automated seizure detection maintained robust accuracy despite drastic spatial down-sampling of recording channels simulating sub-scalp devices. The dominant effects of patient and algorithm on detection accuracy highlight the need for both careful patient selection and continued development of seizure detection algorithms optimized for the unique signal characteristics of sub-scalp and other chronic EEG devices. The correlation between full-montage and reduced-montage performance within patients offers a practical path toward EMU-based patient selection. Together, these findings support reduced-montage sub-scalp devices as a viable approach for ultra-long-term monitoring and advance the methodological foundation needed to bring these devices into routine clinical practice.

## Code and data availability

All code is available at: https://github.com/JoeKojima/Evaluating-Seizure-Detection-Performance-Across-Representative-Reduced-EEG-Montages.git. Intermediate datasets needed to replicate results are available at: https://upenn.box.com/s/o3sfzpmr20t2n1bwa037q00n9jselkt8. Raw EEG data can be made available on reasonable request and with a data use agreement according to IRB requirements.

## Funding

Haoer Shi and Brian Litt received support from the NIH Pioneer Award, DP1-NS-122038, NIH/NINDS R01-NS-125137, NIH T32NS091006, foundation support from Jonathan and Bonnie Rothberg, Neil and Barbara Smit, and the Small Lake Foundation. Will Ojemann received support from the NSF (GRF DGE-1845298). Carlos Aguila received support from the NSF GRFP (GRF DGE-2236662). Juri Kim and Erin Conrad received support from the National Institute of Neurological Disorders and Stroke (NINDS K23 NS121401-01A1) and the Burroughs Wellcome Fund.

## Conflicts of Interest

Haoer Shi has no conflicts to report. Will Ojemann has no conflicts to report. Erin Conrad performed paid consulting work for Epiminder. Taneeta Mindy Ganguly has received research funding and has performed paid consulting work from UNEEG and from Epiminder. Brian Litt serves as a paid consultant for Natus (Scientific Advisory Board), UNEEG (Scientific Advisory Board), EpiMinder (DSMB) and NAMSA. This study was not industry sponsored or supported.

## Notes

### Competing Interest Statement

EC performed paid consulting work for Epiminder. TMG has received research funding and has performed paid consulting work from UNEEG and from Epiminder. BL serves as a paid consultant for Natus (Scientific Advisory Board), UNEEG (Scientific Advisory Board), EpiMinder (DSMB) and NAMSA. Other authors HS have no conflicts to report. This study was not industry sponsored or supported.

### Funding Statement

HS and BL received support from the NIH Pioneer Award, DP1-NS-122038, NIH/NINDS R01-NS-125137, NIH T32NS091006, foundation support from Jonathan and Bonnie Rothberg, Neil and Barbara Smit, and the Small Lake Foundation. WO received support from the NSF (GRF DGE-1845298). CA received support from the NSF GRFP (GRF DGE-2236662). JK and EC received support from the National Institute of Neurological Disorders and Stroke (NINDS K23 NS121401-01A1) and the Burroughs Wellcome Fund.

### Author Declarations

The Institutional Review Board (IRB) of the University of Pennsylvania gave ethical approval for this work.

