## Supplementary Materials for "How Much Does the Reduced EEG Montage Matter for Seizure Detection?: A Large-Cohort Simulation Study"

### Supplementary Online Content

#### eMethods

##### EEG data acquisition
EEG data were acquired using multiple types of amplifiers according to clinical needs, with a sampling rate of 256 Hz. The electrodes were placed according to the international 10-20 system. In some cases, additional subtemporal electrodes were placed, and we excluded these from all analyses. The machine reference electrode was placed behind Cz.

##### Seizure and interictal clip identification
We included consecutive admissions from the Hospital of the University of Pennsylvania (HUP) Epilepsy Monitoring Unit (EMU) from 2017 to 2024. Data from 2017 to 2022 consisted of technologist-clipped segments containing seizures and representative interictal clips, whereas data from 2023 to 2024 consisted of continuous recordings throughout the entire EMU stay. Seizures confirmed by the treating clinical team in the EMU were identified from annotations embedded within EMU recordings. Annotations containing "UEO" (unequivocal onset) or "EEC" (earliest electrographic change) were considered to mark electrographic seizure onset, a standard clinical annotation for our center. When multiple "UEO" or "EEC" annotations were present for a single event, the earliest timestamp was defined as seizure onset. Seizure offset was defined as the first subsequent annotation containing "OFF" or "OFFSET" following the identified onset, also standard terminology for our center. Seizure offset was considered undefined if no "OFF" or "OFFSET" annotation was present within 20 minutes following onset. Interictal clips were derived from non-seizure segments separated by at least 20 minutes from any seizure-related annotation, including "UEO," "EEC," "seizure," "sz," "OFF," or "OFFSET". As only clipped segments were available for data from 2017 to 2022, interictal clips may be unavailable for some admissions and 5-minute pre- and post-ictal padding was sometimes incomplete.

##### Seizure detectors
*SVM*. The SVM model detects seizure as the novelty class based on feature patterns [1]. The EEG data were downsampled to 200 Hz. Three features, including mean curve length, mean energy, and mean Teager energy, were calculated per channel using 1-second windows with 0.5-second overlap. A one-class SVM model with a radial basis function kernel ($\gamma = 1.0$, $\nu = 0.1$) was trained on the first 60 seconds of the earliest interictal clip of each patient. The model follows a novelty detection framework, where $\nu$ specifies the expected fraction of outliers in interictal data; seizure activity is assumed to produce a substantially higher empirical outlier fraction, enabling detection. A sample was labeled as seizure if the proportion of novel samples within the 20 most recent windows exceeded the threshold. A 180-second refractory period was enforced, such that additional detections within this interval were suppressed and treated as a single event.

*SPaRCNet*. SPaRCNet is a deep convolutional neural network for seizure detection, trained on over 6,000 EEG recordings from 2,711 patients and rigorously validated [2]. The model discriminates among six types of ictal-interictal injury continuum patterns. Seizure probability was defined using the lateralized periodic discharges (LPD) class rather than the seizure class, due to empirically higher sensitivity on visual review of a subset of our data. Predictions were generated on 10-second windows with 2-second overlap. When passing reduced montage data, all remaining channels were set to zero due to the requirement of 16-channel input.

*NDD*. NDD (Neural Dynamic Divergence) detects seizure activity by measuring deviation from patient-specific baseline neural dynamics [3]. For each patient, a non-linear long short-term memory (LSTM) model was trained to forecast future brain states from input observations. We trained the model for 10 epochs on a one-minute interictal segment. Seizure activity was then detected when the model loss, which served as the seizure probability scores, elevates beyond the baseline level. Predictions were made on 1-second clips with 0.5-second overlap. The model generated predictions for each channel, and a sample was classified as ictal if the average probability across channels exceeded a predefined threshold.

**Performance evaluation**

To determine the decision threshold for each montage and detector, predicted seizure probabilities across all admissions were pooled to construct a receiver operating characteristic (ROC) curve, and the threshold maximizing the event-level F1 score was selected. This threshold was applied to generate binary predictions, which were smoothed using morphological binary opening and closing operations (SciPy) to remove isolated positive predictions and fill short gaps between consecutive detections. Consecutive positive (seizure) predictions were grouped into seizure events; events separated by <4 seconds were merged, and those shorter than 20 seconds were discarded to reduce false alarms due to transient noise. Following the SzCORE framework [4], seizure events separated by <90 seconds were further merged and events longer than 5 minutes were split into separate events. We also reported the area under the ROC curve (AUROC) and the area under the precision-recall curve (AUPRC) for threshold-independent performances.

**Proportion of variance explained**

To explore the proportion of variance explained by patient (admissions), detector, and montage, a linear mixed-effects model was fitted with F1 score as outcome, detector and montage as categorical fixed effects and their interaction included, and a random intercept for admission to account for repeated measures. For inference on fixed effects, Type III Wald tests of model terms were computed. Model fit was summarized using marginal $R^2$ (variance explained by fixed effects) and conditional $R^2$ (variance explained by fixed and random effects), as described by Nakagawa and Schielzeth [5]. Variance decomposition was performed to estimate the proportion attributable to residual variance, admission-level random effects, and incremental contributions of fixed effects based on changes in marginal $R^2$.

**Linear mixed-effects model on epilepsy characteristics**

To assess the influence of epilepsy characteristics on performance, we fitted three linear mixed-effects models for each detector, examining the effects of epilepsy classification (focal versus generalized), lateralization (left versus right versus bilateral), and localization (temporal versus frontal) separately. Admissions were included as random effects, while montage, the epilepsy characteristic of interest, and their interaction were included as fixed effects. For models for lateralization and localization, the other was additionally controlled as a covariate.

**Performance Concordance**

To assess whether full-montage EMU performance predicts reduced-montage behavior in the same patient, we calculated Spearman correlations between full and reduced F1 scores across admissions.

**Basic feature calculation**

Features were computed on a per-channel basis in non-overlapping 5-second windows for 16 standard bipolar channels in the 10–20 montage. Time-domain features included: (1) variance, defined as the mean squared deviation from the channel mean; (2) amplitude, defined as the mean absolute signal value; (3) envelope, defined as the mean absolute amplitude of the Hilbert-transformed analytic signal; and (4) line length, defined as the mean absolute difference between consecutive samples. Frequency-domain features included bandpower in six canonical bands: delta (1–4 Hz), theta (4–8 Hz), alpha (8–13 Hz), sigma (12–16 Hz), beta (13–30 Hz), and gamma (30–40 Hz). Power spectral density was estimated using Welch's method. For clip-level analyses, features were averaged across windows spanning seizure onset to offset for ictal clips, and across all windows for interictal clips. We then compared feature distributions between missed and detected seizures, as well as between missed seizures and interictal clips.

**Feature importance estimation**

A logistic regression model was fitted with missed status as the outcome and log-transformed, standardized features as predictors. A 5-fold stratified cross-validation at the admission level was used. Feature importance was assessed within each fold using permutation importance on the validation set [6]. Specifically, for each feature, we performed 30 permutations of its validation set values and calculated the mean decrease in area under the receiver operating characteristics curve (AUROC) relative to baseline. Feature importances were averaged across permutations and folds.

#### eTable 1. List of simulated montages and bipolar channels used

| Montage | Laterality | Channels |
| --- | --- | --- |
| Full | Bilateral | Fp1-F7, F7-T3, T3-T5, T5-O1, Fp1-F3, F3-C3, C3-P3, P3-O1, Fp2-F8, F8-T4, T4-T6, T6-O2, Fp2-F4, F4-C4, C4-P4, P4-O2 |
| Centropariental | Bilateral | C3-P3, C4-P4 |
| Frontotemporal | Left | F3-T3 |
| | Right | F4-T4 |
| | Bilateral | F3-T3, F4-T4 |
| Temporal | Left | F7-T3 |
| | Right | F8-T4 |
| | Bilateral | F7-T3, F8-T4 |
| Centrotemporal | Left | C3-T3 |
| | Right | C4-T4 |
| | Bilateral | C3-T3, C4-T4 |
| Temporoparietal | Left | T3-P3 |
| | Right | T4-P4 |
| | Bilateral | T3-P3, T4-P4 |
| Posterior temporal | Left | T3-T5 |
| | Right | T4-T6 |
| | Bilateral | T3-T5, T4-T6 |
| Circumferential | Bilateral | Fp1-F7, F7-T3, T3-T5, T5-O1, Fp2-F8, F8-T4, T4-T6, T6-O2 |

#### eTable 2. Linear mixed effects model results and proportion of variance explained

| | | M0 | | M1 | | M2 | | M3 | |
|---|---|---|---|---|---|---|---|---|---|
| **Term** | **Df** | $\chi^2$ | ***p*** | $\chi^2$ | ***p*** | $\chi^2$ | ***p*** | $\chi^2$ | ***p*** |
| **(Intercept)** | 1 | 4482.4 | <.001 *** | 4228.6 | <.001 *** | 2968.7 | <.001 *** | 2019.7 | <.001 *** |
| **Detector** | 2 | / | / | 1183.7 | <.001 *** | 1192.0 | <.001 *** | 167.3 | <.001 *** |
| **Montage** | 4 | / | / | / | / | 45.49 | <.001 *** | 1.951 | 0.745 |
| **Detector × Montage** | 8 | / | / | / | / | / | / | 63.34 | <.001 *** |
| **Marginal $R^2$** | | 0 | | 0.103 | | 0.107 | | 0.112 | |
| **Conditional $R^2$** | | 0.284 | | 0.394 | | 0.398 | | 0.404 | |
| **Incremental $\Delta R^2$** | | / | | 0.103 (model) | | 0.004 (montage) | | 0.005 (interaction) | |
| **Cond. $\Delta R^2$ - Marg. $R^2$** | | 0.284 | | 0.291 | | 0.291 | | 0.292 (admission) | |

M0: F1 ~ 1 + (1 | admission)

M1: F1 ~ detector + (1 | admission)

M2: F1 ~ detector + montage + (1 | admission)

M3: F1 ~ detector * montage + (1 | admission)

Linear mixed effects models M0-M3 were built following the formula above. Df, degree of freedom; $\chi^2$ , chi-square; chi-square and p-values were obtained through Type III Wald tests. The marginal and conditional $R^2$ were calculated following the Nakagawa and Schielzeth method [5]. The proportion of variance explained by each fixed factor was calculated as the change in marginal $R^2$ between incremental models, while the variance explained by random effects was computed as the difference between the conditional and marginal $R^2$ of the final model (M3). ***, *p*<.001.

#### eTable 3. SPaRCNet seizure detection performance across all simulated montages

| Montage | AUROC | AUPRC | Sensitivity | FA / Hour | F1 Score |
|---|---|---|---|---|---|
| Full | 0.841 [0.827–0.853] | 0.546 [0.518–0.571] | 0.747 [0.713–0.781] | 1.532 [1.351–1.723] | 0.645 [0.615–0.675] |
| C3-P3, C4-P4 | 0.763 [0.748–0.778] | 0.406 [0.380–0.432] | 0.784 [0.751–0.815] | 3.365 [3.089–3.636] | 0.565 [0.538–0.591] |
| F3-T3 | 0.805 [0.791–0.819] | 0.502 [0.479–0.528] | 0.756 [0.724–0.789] | 2.032 [1.802–2.271] | 0.620 [0.591–0.650] |
| F4-T4 | 0.780 [0.763–0.795] | 0.438 [0.412–0.464] | 0.797 [0.767–0.826] | 3.609 [3.319–3.876] | 0.568 [0.541–0.594] |
| F3-T3, F4-T4 | 0.854 [0.841–0.866] | 0.519 [0.493–0.546] | 0.858 [0.830–0.883] | 3.086 [2.830–3.354] | 0.640 [0.615–0.665] |
| F7-T3 | 0.527 [0.508–0.545] | 0.250 [0.231–0.272] | 0.986 [0.976–0.994] | 8.523 [8.397–8.649] | 0.513 [0.500–0.526] |
| F8-T4 | 0.472 [0.453–0.491] | 0.191 [0.174–0.209] | 0.988 [0.981–0.994] | 8.525 [8.402–8.650] | 0.521 [0.508–0.533] |
| F7-T3, F8-T4 | 0.548 [0.527–0.568] | 0.261 [0.238–0.285] | 0.984 [0.975–0.992] | 8.507 [8.379–8.624] | 0.507 [0.494–0.520] |
| C3-T3 | 0.730 [0.715–0.747] | 0.385 [0.363–0.411] | 0.765 [0.735–0.798] | 3.159 [2.903–3.423] | 0.568 [0.543–0.595] |
| C4-T4 | 0.765 [0.749–0.780] | 0.425 [0.400–0.450] | 0.801 [0.771–0.831] | 3.483 [3.209–3.755] | 0.574 [0.549–0.600] |
| C3-T3, C4-T4 | 0.806 [0.792–0.819] | 0.435 [0.410–0.461] | 0.884 [0.860–0.908] | 4.269 [3.992–4.539] | 0.598 [0.576–0.619] |
| P3-T3 | 0.769 [0.755–0.785] | 0.442 [0.418–0.469] | 0.867 [0.841–0.891] | 3.866 [3.589–4.157] | 0.624 [0.601–0.648] |
| P4-T4 | 0.764 [0.749–0.779] | 0.413 [0.390–0.439] | 0.856 [0.828–0.881] | 3.792 [3.535–4.065] | 0.614 [0.588–0.638] |
| P3-T3, P4-T4 | 0.705 [0.688–0.721] | 0.358 [0.336–0.381] | 0.856 [0.829–0.882] | 4.179 [3.906–4.448] | 0.598 [0.574–0.622] |
| T3-T5 | 0.646 [0.628–0.666] | 0.387 [0.364–0.412] | 0.995 [0.990–0.999] | 8.462 [8.349–8.558] | 0.520 [0.509–0.530] |
| T4-T6 | 0.546 [0.527–0.565] | 0.281 [0.260–0.302] | 0.988 [0.980–0.995] | 8.489 [8.371–8.617] | 0.514 [0.502–0.526] |
| T3-T5, T4-T6 | 0.666 [0.648–0.684] | 0.387 [0.363–0.411] | 0.987 [0.979–0.995] | 8.489 [8.356–8.625] | 0.507 [0.495–0.519] |
| Circumferential | 0.714 [0.694–0.731] | 0.407 [0.381–0.432] | 0.664 [0.630–0.698] | 2.644 [2.400–2.912] | 0.515 [0.487–0.544] |

Cell shows mean [95%CI]. AUROC, area under the receiver operating characteristics curve; AUPRC, area under the precision-recall curve; FA/Hour, false alarms per hour.

#### eTable 4. NDD seizure detection performance across all simulated montages

| Montage | AUROC | AUPRC | Sensitivity | FA / Hour | F1 Score |
|---|---|---|---|---|---|
| Full | 0.810 [0.796–0.823] | 0.331 [0.306–0.356] | 0.760 [0.728–0.795] | 2.838 [2.614–3.029] | 0.558 [0.532–0.585] |
| C3-P3, C4-P4 | 0.821 [0.809–0.833] | 0.334 [0.311–0.358] | 0.815 [0.782–0.844] | 3.371 [3.170–3.561] | 0.572 [0.549–0.597] |
| F3-T3 | 0.805 [0.792–0.819] | 0.332 [0.307–0.356] | 0.756 [0.723–0.789] | 2.788 [2.574–2.983] | 0.561 [0.533–0.587] |
| F4-T4 | 0.799 [0.785–0.813] | 0.325 [0.302–0.349] | 0.769 [0.735–0.801] | 2.919 [2.685–3.129] | 0.559 [0.531–0.584] |
| F3-T3, F4-T4 | 0.810 [0.796–0.824] | 0.336 [0.312–0.362] | 0.827 [0.798–0.859] | 3.470 [3.245–3.675] | 0.572 [0.546–0.597] |
| F7-T3 | 0.790 [0.776–0.805] | 0.314 [0.291–0.338] | 0.802 [0.769–0.833] | 3.432 [3.204–3.656] | 0.554 [0.529–0.580] |
| F8-T4 | 0.787 [0.773–0.801] | 0.308 [0.286–0.331] | 0.731 [0.698–0.765] | 2.772 [2.552–2.976] | 0.532 [0.505–0.560] |
| F7-T3, F8-T4 | 0.797 [0.784–0.813] | 0.323 [0.299–0.346] | 0.739 [0.704–0.772] | 2.684 [2.480–2.891] | 0.544 [0.516–0.570] |
| C3-T3 | 0.811 [0.798–0.824] | 0.325 [0.303–0.349] | 0.788 [0.757–0.819] | 2.990 [2.758–3.189] | 0.573 [0.548–0.600] |
| C4-T4 | 0.809 [0.795–0.822] | 0.326 [0.304–0.349] | 0.808 [0.779–0.839] | 3.501 [3.270–3.720] | 0.557 [0.533–0.581] |
| C3-T3, C4-T4 | 0.820 [0.807–0.833] | 0.336 [0.313–0.360] | 0.822 [0.792–0.851] | 3.493 [3.262–3.726] | 0.572 [0.547–0.597] |
| P3-T3 | 0.814 [0.801–0.827] | 0.330 [0.308–0.355] | 0.802 [0.772–0.833] | 3.338 [3.119–3.545] | 0.568 [0.544–0.593] |
| P4-T4 | 0.809 [0.796–0.823] | 0.329 [0.308–0.353] | 0.822 [0.793–0.852] | 3.722 [3.492–3.944] | 0.548 [0.523–0.571] |
| P3-T3, P4-T4 | 0.822 [0.809–0.835] | 0.341 [0.319–0.366] | 0.836 [0.807–0.865] | 3.733 [3.505–3.935] | 0.562 [0.537–0.585] |
| T3-T5 | 0.810 [0.797–0.825] | 0.337 [0.314–0.362] | 0.757 [0.725–0.791] | 2.786 [2.584–2.984] | 0.559 [0.534–0.586] |
| T4-T6 | 0.802 [0.788–0.817] | 0.329 [0.307–0.354] | 0.777 [0.746–0.808] | 3.185 [2.966–3.381] | 0.546 [0.521–0.570] |
| T3-T5, T4-T6 | 0.816 [0.803–0.830] | 0.343 [0.320–0.369] | 0.764 [0.731–0.796] | 2.850 [2.632–3.054] | 0.560 [0.535–0.586] |
| Circumferential | 0.810 [0.796–0.824] | 0.335 [0.310–0.360] | 0.733 [0.699–0.770] | 2.512 [2.313–2.708] | 0.554 [0.528–0.584] |

Cell shows mean [95%CI]. AUROC, area under the receiver operating characteristics curve; AUPRC, area under the precision-recall curve; FA/Hour, false alarms per hour.

#### eTable 5. SVM seizure detection performance across all simulated montages

| Montage | AUROC | AUPRC | Sensitivity | FA / Hour | F1 Score |
|---|---|---|---|---|---|
| Full | 0.708 [0.691–0.726] | 0.228 [0.207–0.248] | 0.649 [0.613–0.684] | 3.734 [3.516–3.951] | 0.452 [0.426–0.479] |
| C3-P3, C4-P4 | 0.616 [0.596–0.636] | 0.150 [0.137–0.165] | 0.671 [0.639–0.705] | 5.482 [5.266–5.710] | 0.404 [0.382–0.426] |
| F3-T3 | 0.558 [0.539–0.577] | 0.104 [0.095–0.113] | 0.656 [0.623–0.692] | 6.450 [6.229–6.678] | 0.368 [0.348–0.391] |
| F4-T4 | 0.552 [0.534–0.570] | 0.101 [0.092–0.110] | 0.666 [0.632–0.698] | 6.665 [6.444–6.887] | 0.362 [0.343–0.382] |
| F3-T3, F4-T4 | 0.568 [0.549–0.587] | 0.118 [0.108–0.130] | 0.696 [0.663–0.728] | 6.977 [6.768–7.173] | 0.369 [0.351–0.388] |
| F7-T3 | 0.525 [0.507–0.544] | 0.089 [0.083–0.097] | 0.732 [0.701–0.761] | 8.075 [7.887–8.254] | 0.363 [0.344–0.380] |
| F8-T4 | 0.529 [0.512–0.547] | 0.091 [0.084–0.099] | 0.683 [0.650–0.715] | 7.172 [6.953–7.369] | 0.359 [0.340–0.378] |
| F7-T3, F8-T4 | 0.536 [0.517–0.557] | 0.102 [0.093–0.112] | 0.671 [0.638–0.707] | 6.622 [6.398–6.834] | 0.367 [0.347–0.388] |
| C3-T3 | 0.575 [0.556–0.594] | 0.106 [0.097–0.115] | 0.652 [0.617–0.689] | 6.064 [5.819–6.281] | 0.374 [0.353–0.396] |
| C4-T4 | 0.574 [0.557–0.592] | 0.104 [0.095–0.114] | 0.688 [0.658–0.722] | 6.806 [6.579–7.020] | 0.374 [0.355–0.392] |
| C3-T3, C4-T4 | 0.590 [0.571–0.609] | 0.123 [0.113–0.135] | 0.712 [0.680–0.743] | 6.848 [6.637–7.054] | 0.383 [0.365–0.401] |
| P3-T3 | 0.576 [0.557–0.595] | 0.106 [0.097–0.116] | 0.726 [0.695–0.757] | 7.225 [7.030–7.440] | 0.376 [0.358–0.394] |
| P4-T4 | 0.570 [0.552–0.587] | 0.106 [0.097–0.116] | 0.708 [0.681–0.739] | 6.941 [6.723–7.163] | 0.383 [0.365–0.401] |
| P3-T3, P4-T4 | 0.588 [0.570–0.608] | 0.123 [0.111–0.134] | 0.704 [0.673–0.737] | 6.898 [6.691–7.109] | 0.378 [0.359–0.397] |
| T3-T5 | 0.552 [0.533–0.569] | 0.097 [0.088–0.105] | 0.721 [0.688–0.753] | 7.277 [7.074–7.479] | 0.381 [0.362–0.399] |
| T4-T6 | 0.551 [0.534–0.568] | 0.095 [0.087–0.103] | 0.708 [0.675–0.737] | 7.656 [7.467–7.854] | 0.365 [0.346–0.383] |
| T3-T5, T4-T6 | 0.564 [0.545–0.584] | 0.110 [0.100–0.120] | 0.708 [0.678–0.742] | 6.994 [6.795–7.185] | 0.375 [0.358–0.395] |
| Circumferential | 0.580 [0.560–0.601] | 0.147 [0.133–0.162] | 0.662 [0.627–0.696] | 6.023 [5.804–6.256] | 0.380 [0.359–0.400] |

Cell shows mean [95%CI]. AUROC, area under the receiver operating characteristics curve; AUPRC, area under the precision-recall curve; FA/Hour, false alarms per hour.

#### eTable 6. Linear mixed effects model results on montage and epilepsy characteristics

| | Model | SPaRCNet | | NDD | |
|---|---|---|---|---|---|
| **Term** | **Df** | $\chi^2$ | ***p*** | $\chi^2$ | ***p*** |
| **Epilepsy classification (n=401)** | | | | | |
| **(Intercept)** | 1 | 1343.0 | <.001 *** | 1444.4 | <.001 *** |
| **Montage** | 4 | 53.33 | <.001 *** | 6.074 | 0.194 |
| **Classification** | 1 | 1.237 | 0.266 | 5.001 | 0.025 * |
| **Montage × Classification** | 4 | 3.288 | 0.511 | 8.663 | 0.070 |
| **Epilepsy lateralization (n=340)** | | | | | |
| **(Intercept)** | 1 | 115.1 | <.001 *** | 99.98 | <.001 *** |
| **Montage** | 4 | 2.416 | 0.660 | 9.229 | 0.056 |
| **Lateralization** | 2 | 8.113 | 0.017 * | 0.956 | 0.620 |
| **Localization** | 3 | 6.009 | 0.111 | 0.875 | 0.831 |
| **Montage × Lateralization** | 8 | 30.21 | <.001 *** | 12.05 | 0.149 |
| **Epilepsy localization (n=285)** | | | | | |
| **(Intercept)** | 1 | 89.77 | <.001 *** | 101.7 | <.001 *** |
| **Montage** | 4 | 7.796 | 0.099 . | 7.939 | 0.094 |
| **Localization** | 1 | 0.524 | 0.469 | 3.333 | 0.068 |
| **Lateralization** | 3 | 1.470 | 0.689 | 0.763 | 0.858 |
| **Montage × Localization** | 4 | 9.021 | 0.061 . | 6.608 | 0.158 |

Three linear mixed-effects models were built for each detector (SPaRCNet and NDD), examining the effects of epilepsy classification (focal versus generalized, n=401), lateralization (left versus right versus bilateral, n=340), and localization (temporal versus frontal, n=285) separately. Admissions were included as random effects, while montage, the epilepsy characteristic of interest, and their interaction were included as fixed effects. For models for lateralization and localization, the other was additionally controlled as a covariate. Df, degree of freedom; $\chi^2$ , chi-square; chi-square and p-values were obtained through Type III Wald tests.

**eFigure 1. Flow diagram of data cohort**

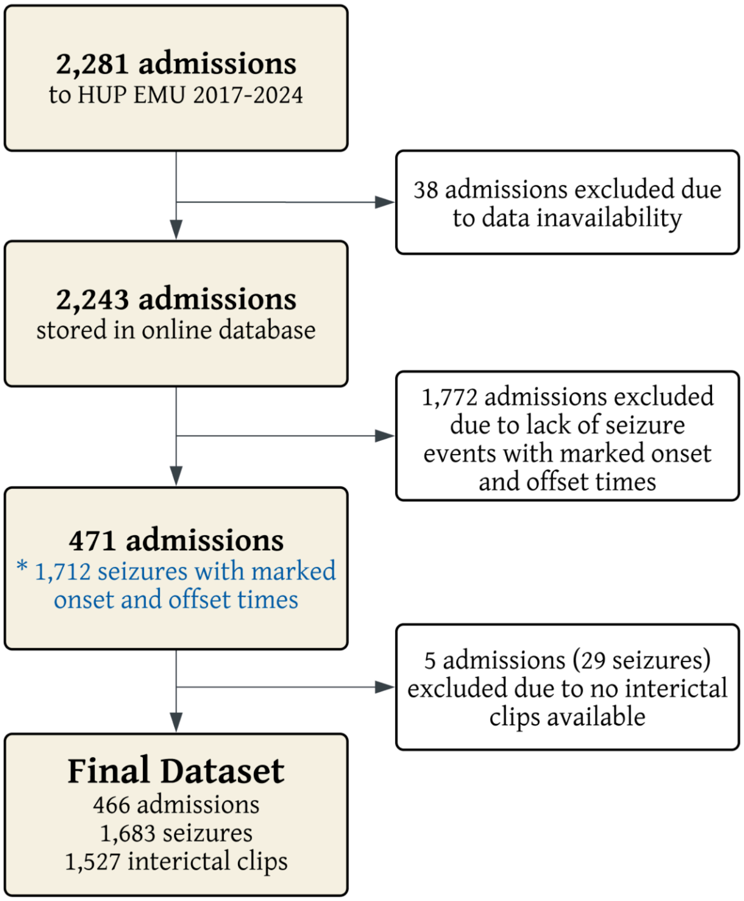

**eFigure 2. Receiver-operating curves and precision-recall curves across models and montages**

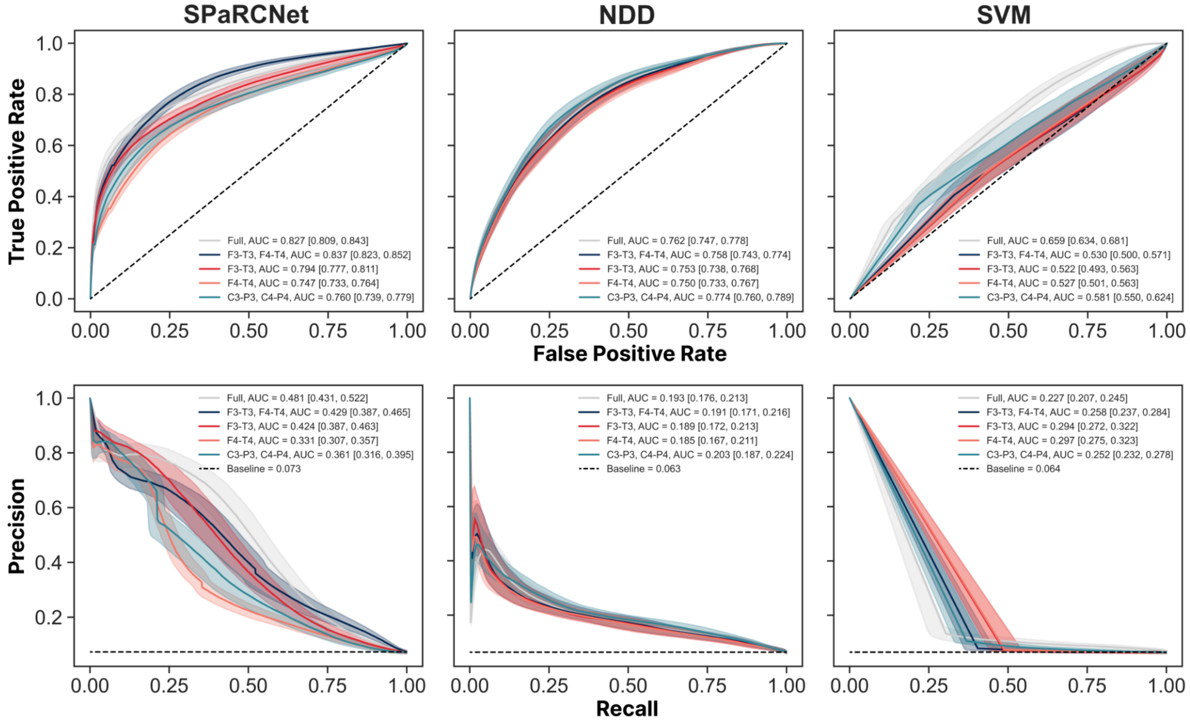


* Shaded areas and numbers in brackets represent 95% CIs from 1000 admission-level bootstraps. AUC, area under curve.

#### eFigure 3: Correlation of seizure detection performance across montages

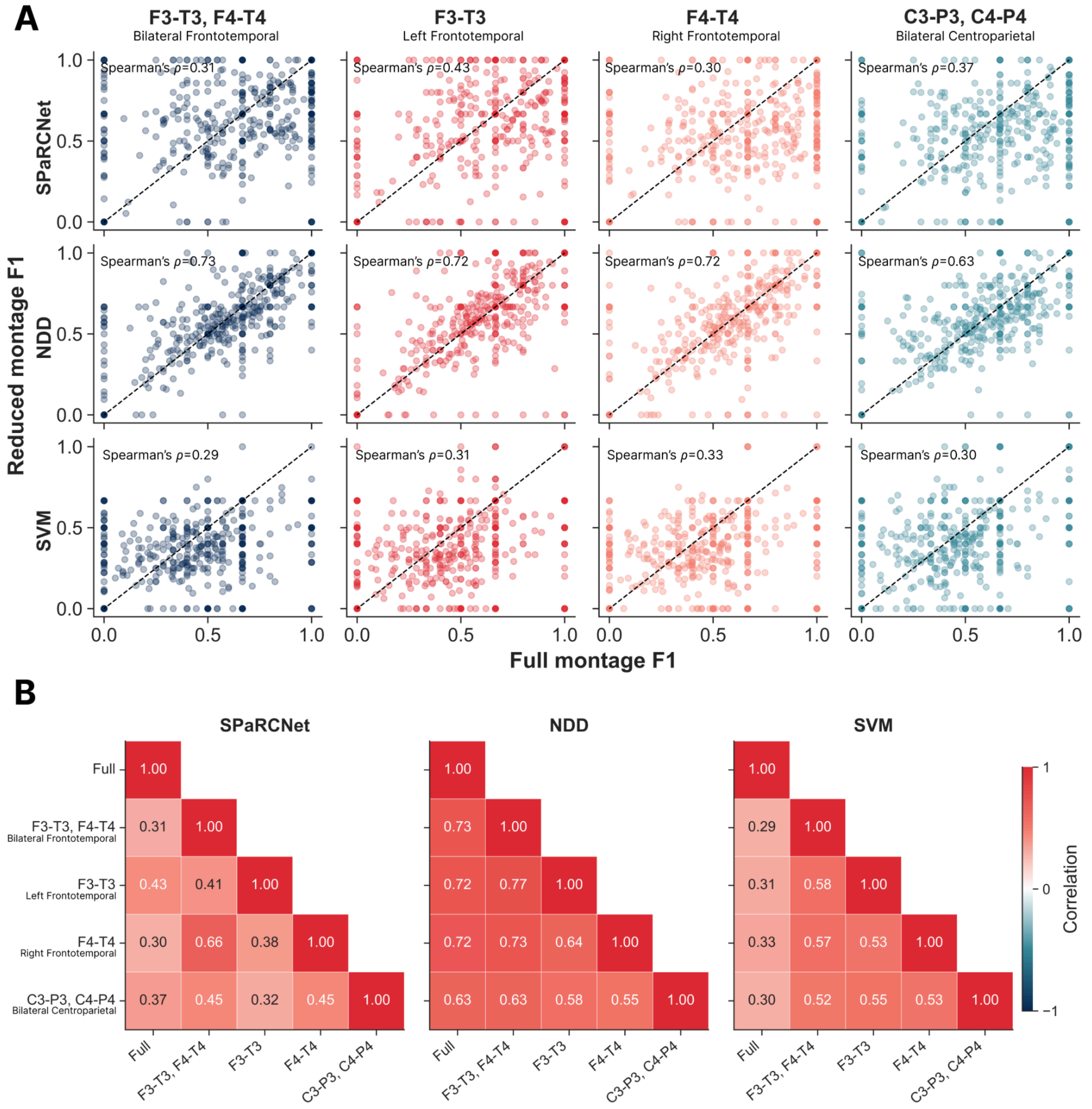


| SPaRCNet | Full | F3-T3, F4-T4 | F3-T3 | F4-T4 | C3-P3, C4-P4 |
|---|---|---|---|---|---|
| Full | 1.00 | | | | |
| F3-T3, F4-T4 | 0.31 | 1.00 | | | |
| F3-T3 | 0.43 | 0.41 | 1.00 | | |
| F4-T4 | 0.30 | 0.66 | 0.38 | 1.00 | |
| C3-P3, C4-P4 | 0.37 | 0.45 | 0.32 | 0.45 | 1.00 |

| NDD | Full | F3-T3, F4-T4 | F3-T3 | F4-T4 | C3-P3, C4-P4 |
|---|---|---|---|---|---|
| Full | 1.00 | | | | |
| F3-T3, F4-T4 | 0.73 | 1.00 | | | |
| F3-T3 | 0.72 | 0.77 | 1.00 | | |
| F4-T4 | 0.72 | 0.73 | 0.64 | 1.00 | |
| C3-P3, C4-P4 | 0.63 | 0.63 | 0.58 | 0.55 | 1.00 |

| SVM | Full | F3-T3, F4-T4 | F3-T3 | F4-T4 | C3-P3, C4-P4 |
|---|---|---|---|---|---|
| Full | 1.00 | | | | |
| F3-T3, F4-T4 | 0.29 | 1.00 | | | |
| F3-T3 | 0.31 | 0.58 | 1.00 | | |
| F4-T4 | 0.33 | 0.57 | 0.53 | 1.00 | |
| C3-P3, C4-P4 | 0.30 | 0.52 | 0.55 | 0.53 | 1.00 |

**A**. Scatter plots of full montage F1 scores (horizontal axis) and reduced montage F1 scores (vertical axis) across detectors and montages. Each dot is a single patient admission. **B.** Spearman correlation coefficients between F1 scores evaluated on the same admissions under different montage configurations for each detector (SPaRCNet, NDD, SVM). Color indicates correlation from −1 (blue) through 0 (white) to +1 (red).

#### eFigure 4: Seizure detection performance across clinical subgroups in all simulated montages.

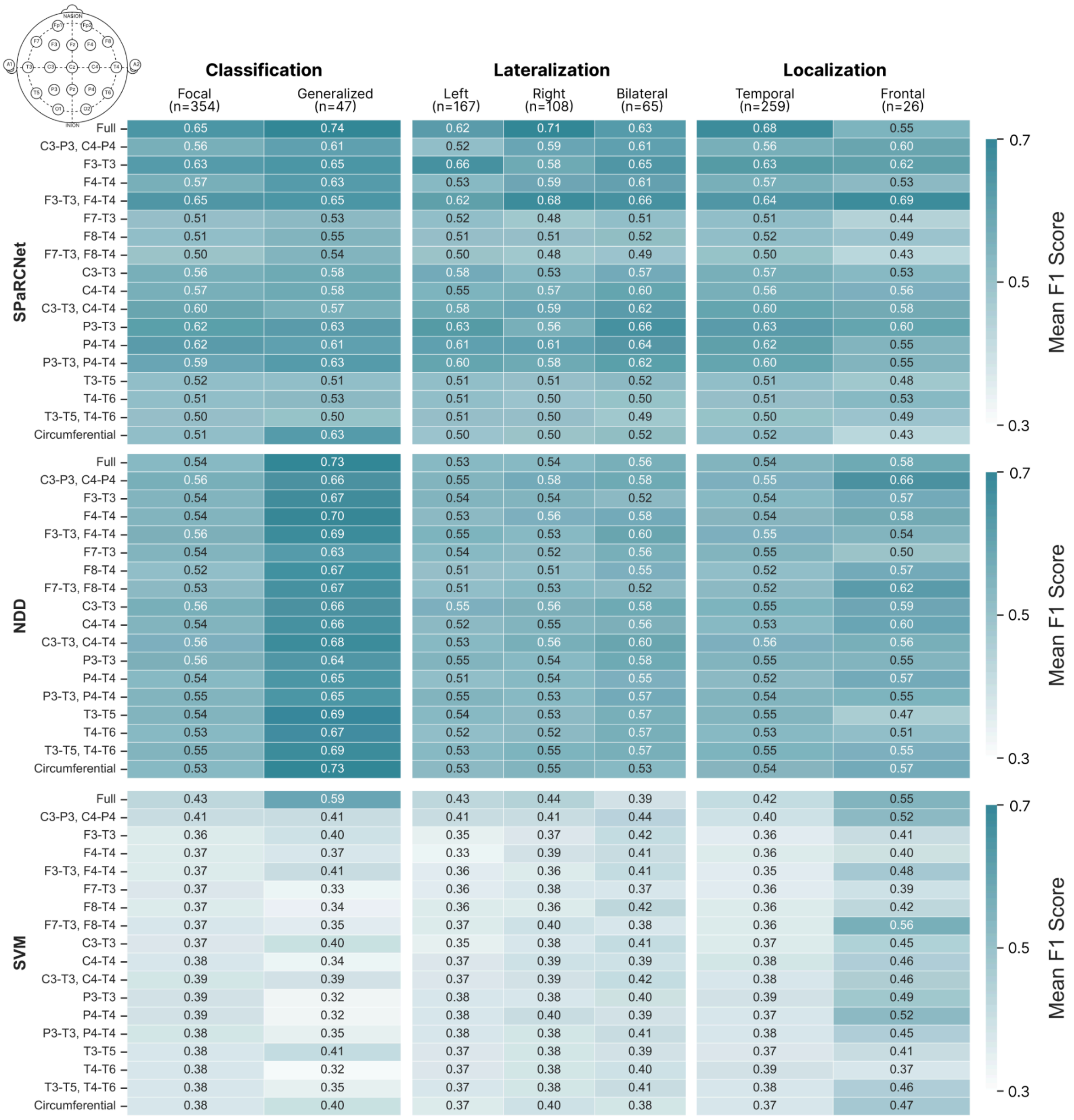


Mean event-wise F1-scores are shown for SPaRCNet (top), NDD (middle), and SVM (bottom) detectors across different electrode montage configurations. Performance is stratified by epilepsy classification (focal, generalized, mixed), laterality (left, right, bilateral), and seizure location (temporal, frontal, multifocal). Each cell represents the mean F1-score across EMU admissions within the corresponding subgroup. Sample sizes were indicated in parentheses. White-to-blue color scale reflects lower to higher performance.
